# Comparative effectiveness of sotrovimab and molnupiravir for prevention of severe COVID-19 outcomes in non-hospitalised patients: an observational cohort study using the OpenSAFELY platform

**DOI:** 10.1101/2022.05.22.22275417

**Authors:** Bang Zheng, Amelia CA Green, John Tazare, Helen J Curtis, Louis Fisher, Linda Nab, Anna Schultze, Viyaasan Mahalingasivam, Edward PK Parker, William J Hulme, Sebastian CJ Bacon, Nicholas J DeVito, Christopher Bates, David Evans, Peter Inglesby, Henry Drysdale, Simon Davy, Jonathan Cockburn, Caroline E Morton, George Hickman, Tom Ward, Rebecca M Smith, John Parry, Frank Hester, Sam Harper, Amir Mehrkar, Rosalind M Eggo, Alex J Walker, Stephen JW Evans, Ian J Douglas, Brian MacKenna, Ben Goldacre, Laurie A Tomlinson

## Abstract

**Objective:** To compare the effectiveness of sotrovimab (a neutralising monoclonal antibody) vs. molnupiravir (an antiviral) in preventing severe COVID-19 outcomes in non-hospitalised high-risk COVID-19 adult patients.

**Design:** With the approval of NHS England, we conducted a real-world cohort study using the OpenSAFELY-TPP platform.

**Setting:** Patient-level electronic health record data were obtained from 24 million people registered with a general practice in England that uses TPP software. The primary care data were securely linked with data on COVID-19 infection and therapeutics, hospital admission, and death within the OpenSAFELY-TPP platform, covering a period where both medications were frequently prescribed in community settings.

**Participants:** Non-hospitalised adult COVID-19 patients at high risk of severe outcomes treated with sotrovimab or molnupiravir since December 16, 2021.

**Interventions:** Sotrovimab or molnupiravir administered in the community by COVID-19 Medicine Delivery Units.

**Main outcome measure:** COVID-19 related hospitalisation or COVID-19 related death within 28 days after treatment initiation.

**Results:** Between December 16, 2021 and February 10, 2022, 3331 and 2689 patients were treated with sotrovimab and molnupiravir, with no substantial differences in their baseline characteristics. The mean age of all 6020 patients was 52 (SD=16) years; 59% were female, 89% White and 88% had three or more COVID-19 vaccinations. Within 28 days after treatment initiation, 87 (1.4%) COVID-19 related hospitalisations/deaths were observed (32 treated with sotrovimab and 55 with molnupiravir). Cox proportional hazards models stratified by area showed that after adjusting for demographics, high-risk cohort categories, vaccination status, calendar time, body mass index and other comorbidities, treatment with sotrovimab was associated with a substantially lower risk than treatment with molnupiravir (hazard ratio, HR=0.54, 95% CI: 0.33 to 0.88; P=0.014). Consistent results were obtained from propensity score weighted Cox models (HR=0.50, 95% CI: 0.31 to 0.81; P=0.005) and when restricted to fully vaccinated people (HR=0.53, 95% CI: 0.31 to 0.90; P=0.019). No substantial effect modifications by other characteristics were detected (all P values for interaction>0.10). Findings were similar in an exploratory analysis of patients treated between February 16 and May 1, 2022 when the Omicron BA.2 variant was dominant in England.

**Conclusion:** In routine care of non-hospitalised high-risk adult patients with COVID-19 in England, those who received sotrovimab were at lower risk of severe COVID-19 outcomes than those receiving molnupiravir.

## Introduction

Neutralising monoclonal antibodies (nMAbs) and antiviral medicines were approved by the UK Medicines and Healthcare products Regulatory Agency (MHRA) for use in non-hospitalised COVID-19 patients to prevent disease progression. On December 16, 2021, COVID-19 Medicine Delivery Units (CMDUs) were launched across England to provide nMAbs and antivirals in community settings to treat symptomatic COVID-19 patients who are at high risk of severe outcomes.

Among the initial available therapeutic options were sotrovimab (an intravenous nMAb) and molnupiravir (an oral antiviral).[1-3] The approval and early clinical use of these medications was based on data from two phase 3 randomised controlled trials (RCTs).[4,5] However, findings of these trials may be limited by relatively small sample size, lack of population generalisability given strict inclusion and exclusion criteria, and the circulating variants prevalent when the trials were conducted. In particular, little evidence is available on their effectiveness in vaccinated COVID-19 patients, patients infected by Omicron variants, or in those with severe renal or liver impairment. There is uncertainty about the efficacy of molnupiravir in patients with prior SARS-CoV-2 infection, diabetes and non-White ethnicity,[5] and the appropriateness of early regulatory authorisation for this drug has been debated given the modest effect magnitude observed in the RCT.[6] In addition, both lower concordance with courses of oral medication and longer time to administration of treatments after symptom onset in routine care compared to clinical trials may impact potential benefit. Therefore, validating the effectiveness of sotrovimab and molnupiravir in preventing adverse outcomes in real-world settings with varied populations is crucial in supporting their wide-scale clinical use among patients with COVID-19.

In the first two months after the launch of CMDUs, sotrovimab and molnupiravir were the most frequently prescribed medications,[2] with anecdotal reports that choice of drug was in part determined by the availability of facilities to deliver intravenous infusions, and relative clinical equipoise for the choice of drug.[3] This provided the opportunity for observational comparison of the effectiveness of the two medications, possibly with limited bias according to patient characteristics. A comparative effectiveness study would also provide real-world evidence for the clinical practice guideline regarding the prioritised treatment.[7]

Therefore, we sought to compare the effectiveness of sotrovimab vs. molnupiravir in preventing severe outcomes from COVID-19 infection among non-hospitalised high-risk adult patients across England during the first two months of the national rollout, utilising near real-time electronic health record (EHR) data in the OpenSAFELY-TPP platform. We also explored the potential modifying effects of different demographic and clinical factors on drug effectiveness. In addition, we conducted an exploratory analysis of patients treated in the following three months when Omicron BA.2 had replaced BA.1 as the dominant variant in England.[8]

## Methods

### Study design and population

In this observational cohort study, we included adult patients (≥18 years old) within the OpenSAFELY-TPP platform who had non-hospitalised treatment records for either sotrovimab or molnupiravir since December 16, 2021 in the COVID-19 therapeutics dataset [2] (**Figure 1**). Our main analyses focused on those treated before February 10, 2022 (Period 1) as after this date treatment recommendations were changed, with molnupiravir moved to third-line.[7] We required patients to be registered at a GP surgery at the time of treatment initiation to allow for extraction of baseline and follow-up information. According to the eligibility criteria from NHS England,[3] to have received COVID-19 nMAb or antiviral treatment in the community during this period, patients needed to have SARS-CoV-2 infection confirmed by polymerase chain reaction (PCR) testing, onset of COVID-19 symptoms within the last five days, and be a member of at least one of the following ten high-risk cohorts: patients with Down syndrome, a solid cancer, a haematological disease or stem cell transplant, renal disease, liver disease, immune-mediated inflammatory disorders, primary immune deficiencies, HIV/AIDS, solid organ transplant, or rare neurological conditions. Supplemental data from patients treated between February 16 (approximate date when the prevalence of Omicron BA.2 variant in England was over 50% [8]) and May 1, 2022 were included in the exploratory analysis (Period 2).

**Figure 1.**
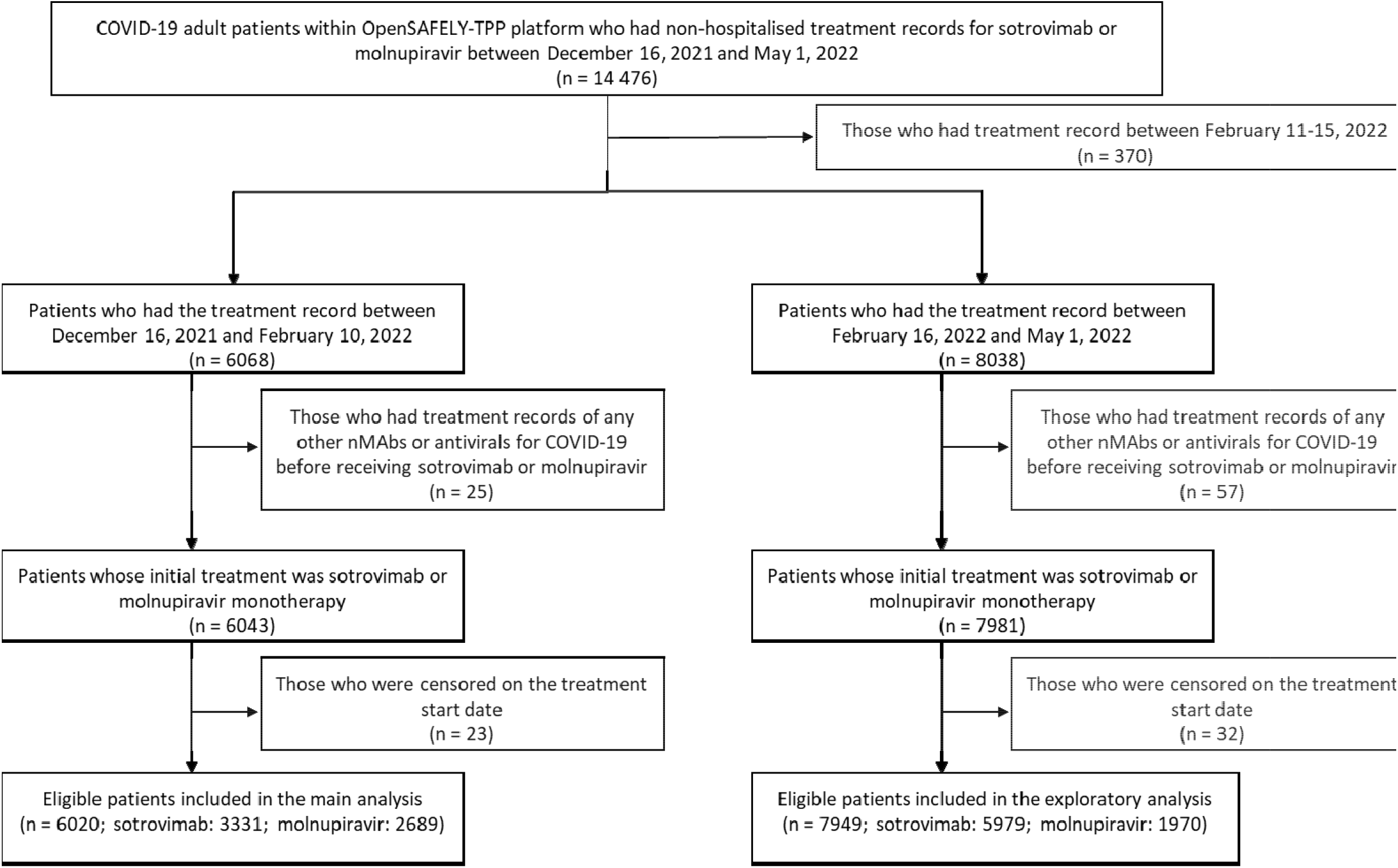
Study population flowchart. Note: nMAbs=neutralising monoclonal antibodies. We included patients treated before May 1, 2022 to allow sufficient follow-up time and time for the linkage data to be updated in OpenSAFELY-TPP platform.

### Data sources

All data were linked, stored and analysed securely within the OpenSAFELY platform: https://opensafely.org/. OpenSAFELY is a data analytics platform created by our team on behalf of NHS England to address urgent COVID-19 research questions. The dataset analysed within OpenSAFELY-TPP is based on 24 million people currently registered with GP surgeries using TPP SystmOne software. Data include pseudonymised data such as coded diagnoses, medications and physiological parameters. No free text data are included. All code is shared openly for review and re-use under MIT open license (https://github.com/opensafely/sotrovimab-and-molnupiravir). Detailed pseudonymised patient data is potentially re-identifiable and therefore not shared. Primary care records managed by the GP software provider TPP are securely linked to other similarly pseudonymised datasets, including the Office for National Statistics (ONS) mortality database, in-patient hospital spell records via Secondary Uses Service (SUS), national coronavirus testing records via the Second Generation Surveillance System (SGSS), and the above-mentioned COVID-19 therapeutics dataset, a patient-level dataset on nMAbs and antiviral treatments derived from Blueteq software that CMDUs use to notify NHS England of COVID-19 treatments. Patient-level vaccination status is available in the GP records directly via the National Immunisation Management System (NIMS).

### Exposure

The exposure of interest was treatment with sotrovimab or molnupiravir administered by CMDUs with date of treatment initiation for each patient as recorded in the COVID-19 therapeutics dataset. Patients were excluded if they had treatment records of any other nMAbs or antivirals for COVID-19 before receiving sotrovimab or molnupiravir (n=25 in Period 1 and 57 in Period 2). Patients with treatment records of both sotrovimab and molnupiravir were censored at the start date of the second treatment (n=10 in Period 1 and 25 in Period 2).

### Outcomes

The primary outcome was COVID-19 related hospitalisation (based on primary diagnosis ascertained from SUS) or COVID-19 related death (based on underlying/contributing causes) within 28 days after treatment initiation. Secondary outcomes were 28-day all-cause hospital admission or death, and 60-day COVID-19 related hospitalisation/death. To exclude events where patients were admitted in order to receive sotrovimab or other planned/regular treatment (e.g., chemotherapy or dialysis), we did not count admissions coded as “elective day case admission” or “regular admission” in SUS or day cases detected by the same admission and discharge dates as hospitalisation events (see **Supplementary Table 1** for breakdown).

**Table 1.**
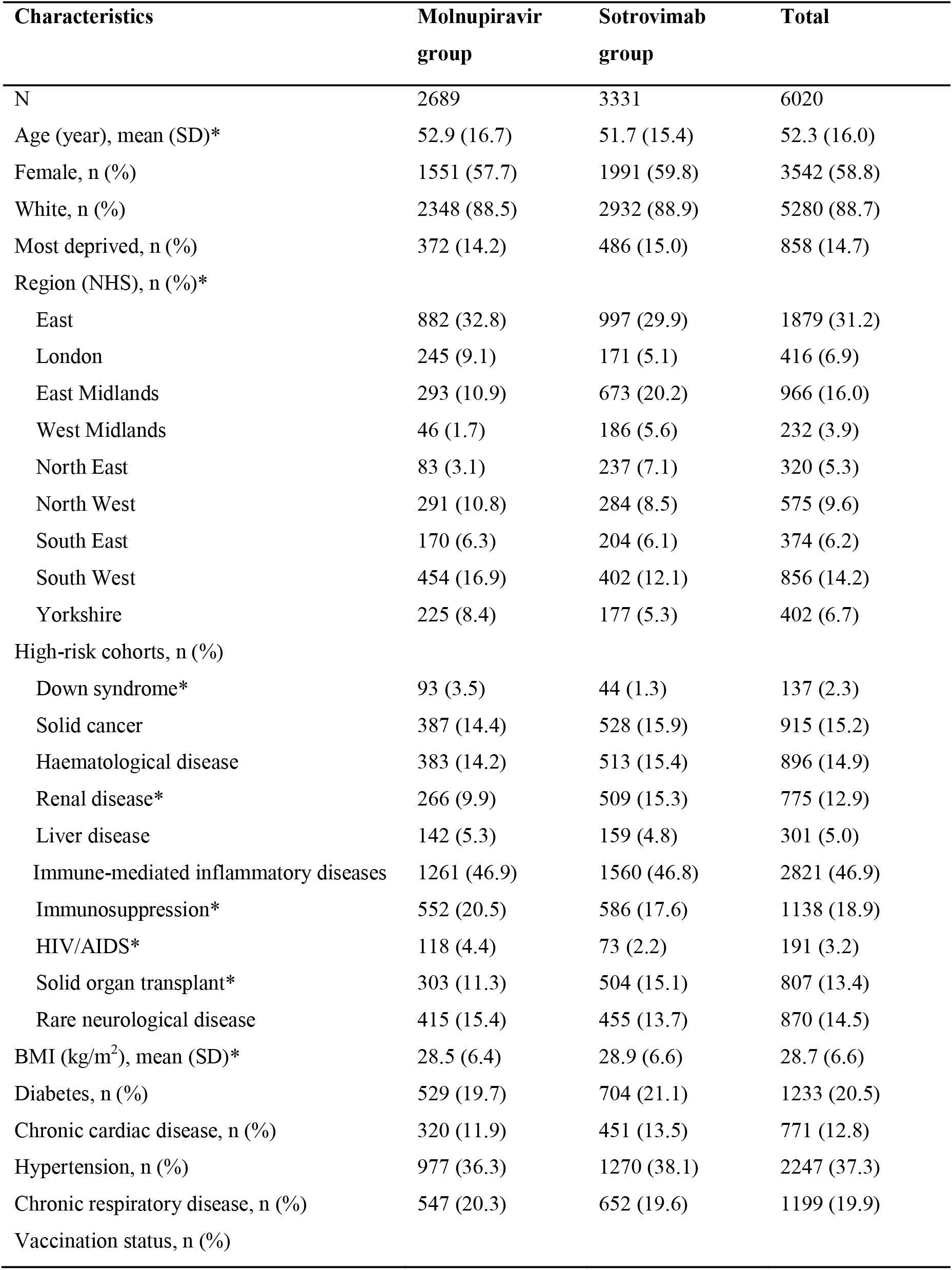
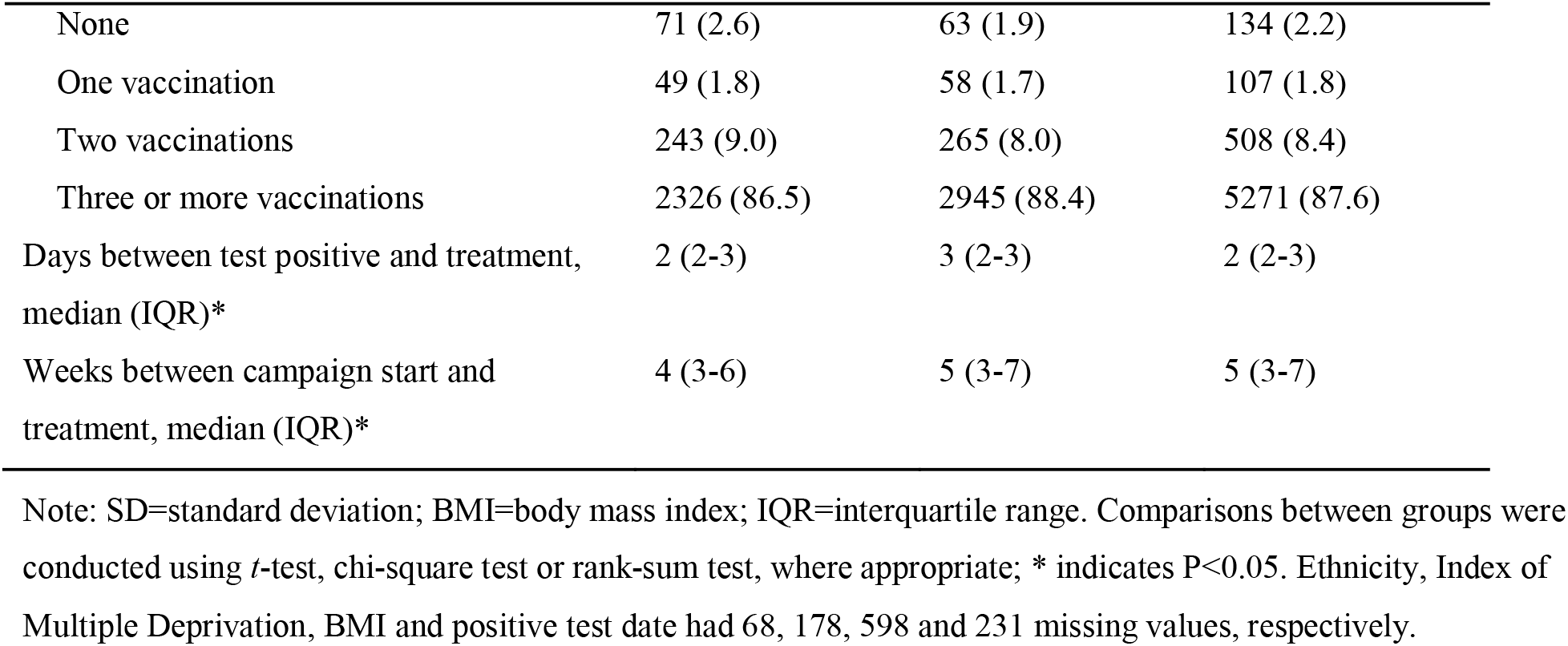
Baseline characteristics of COVID-19 patients receiving molnupiravir or sotrovimab.

| Characteristics | Molnupiravir<br>group | Sotrovimab<br>group | Total |
| --- | --- | --- | --- |
| N | 2689 | 3331 | 6020 |
| Age (year), mean (SD)* | 52.9 (16.7) | 51.7 (15.4) | 52.3 (16.0) |
| Female, n (%) | 1551 (57.7) | 1991 (59.8) | 3542 (58.8) |
| White, n (%) | 2348 (88.5) | 2932 (88.9) | 5280 (88.7) |
| Most deprived, n (%) | 372 (14.2) | 486 (15.0) | 858 (14.7) |
| Region (NHS), n (%)* |  |  |  |
| East | 882 (32.8) | 997 (29.9) | 1879 (31.2) |
| London | 245 (9.1) | 171 (5.1) | 416 (6.9) |
| East Midlands | 293 (10.9) | 673 (20.2) | 966 (16.0) |
| West Midlands | 46 (1.7) | 186 (5.6) | 232 (3.9) |
| North East | 83 (3.1) | 237 (7.1) | 320 (5.3) |
| North West | 291 (10.8) | 284 (8.5) | 575 (9.6) |
| South East | 170 (6.3) | 204 (6.1) | 374 (6.2) |
| South West | 454 (16.9) | 402 (12.1) | 856 (14.2) |
| Yorkshire | 225 (8.4) | 177 (5.3) | 402 (6.7) |
| High-risk cohorts, n (%) |  |  |  |
| Down syndrome* | 93 (3.5) | 44 (1.3) | 137 (2.3) |
| Solid cancer | 387 (14.4) | 528 (15.9) | 915 (15.2) |
| Haematological disease | 383 (14.2) | 513 (15.4) | 896 (14.9) |
| Renal disease* | 266 (9.9) | 509 (15.3) | 775 (12.9) |
| Liver disease | 142 (5.3) | 159 (4.8) | 301 (5.0) |
| Immune-mediated inflammatory diseases | 1261 (46.9) | 1560 (46.8) | 2821 (46.9) |
| Immunosuppression* | 552 (20.5) | 586 (17.6) | 1138 (18.9) |
| HIV/AIDS* | 118 (4.4) | 73 (2.2) | 191 (3.2) |
| Solid organ transplant* | 303 (11.3) | 504 (15.1) | 807 (13.4) |
| Rare neurological disease | 415 (15.4) | 455 (13.7) | 870 (14.5) |
| BMI (kg/m<sup>2</sup>), mean (SD)* | 28.5 (6.4) | 28.9 (6.6) | 28.7 (6.6) |
| Diabetes, n (%) | 529 (19.7) | 704 (21.1) | 1233 (20.5) |
| Chronic cardiac disease, n (%) | 320 (11.9) | 451 (13.5) | 771 (12.8) |
| Hypertension, n (%) | 977 (36.3) | 1270 (38.1) | 2247 (37.3) |
| Chronic respiratory disease, n (%) | 547 (20.3) | 652 (19.6) | 1199 (19.9) |
| Vaccination status, n (%) |  |  |  |

| **Characteristics** | **Molnupiravir**<br>**group** | **Sotrovimab**<br>**group** | **Total** |
| --- | --- | --- | --- |
| None | 71 (2.6) | 63 (1.9) | 134 (2.2) |
| One vaccination | 49 (1.8) | 58 (1.7) | 107 (1.8) |
| Two vaccinations | 243 (9.0) | 265 (8.0) | 508 (8.4) |
| Three or more vaccinations | 2326 (86.5) | 2945 (88.4) | 5271 (87.6) |
| Days between test positive and treatment,<br>median (IQR)* | 2 (2-3) | 3 (2-3) | 2 (2-3) |
| Weeks between campaign start and<br>treatment, median (IQR)* | 4 (3-6) | 5 (3-7) | 5 (3-7) |

Note: SD=standard deviation; BMI=body mass index; IQR=interquartile range. Comparisons between groups were conducted using *t*-test, chi-square test or rank-sum test, where appropriate; \* indicates $P < 0.05$ . Ethnicity, Index of Multiple Deprivation, BMI and positive test date had 68, 178, 598 and 231 missing values, respectively.

### Covariates

The following potential confounding factors or effect modifiers were extracted at baseline: age, sex, Sustainability Transformation Partnerships code of their registered GP surgery (STP, an NHS administrative region assumed to be a proxy for CMDUs), ethnicity (grouped into five broad categories: White, Black or Black British, Asian or Asian British, Mixed, Other), Index of Multiple Deprivation (IMD, as quintiles derived from the patient’s postcode at lower super output area level), calendar week (to account for secular trend of prescription and incidence rate of COVID-19 outcomes), COVID-19 vaccination status (unvaccinated, one vaccination, two vaccinations, or three or more vaccinations), positive test date for SARS-CoV-2 infection (PCR or lateral flow test), body mass index (BMI, the most recent record within 10 years; <18.5 kg/m^2^, 18.5 - <25 kg/m^2^, 25 - <30 kg/m^2^, ≥30 kg/m^2^), ten high-risk cohort categories as mentioned above (allowing multiple categories per patient), and other comorbidities (diabetes, hypertension, chronic cardiac disease and chronic respiratory disease).

Individuals with missing ethnicity, IMD, BMI or positive SARS-CoV-2 test information were included as “Unknown” category for each variable.

### Statistical analyses

Distributions of baseline characteristics were compared between patients treated with sotrovimab vs. molnupiravir using *t*-test, chi-square test or rank-sum test, where appropriate. Follow-up time of individual patients was calculated from the date of the treatment initiation record, until the outcome event date, 28 days after treatment initiation, initiation of a second nMAb/antiviral treatment, death, patient deregistration date, or the study end date (August 10, 2022), whichever occurred first.

Risks of 28-day COVID-19 related hospitalisation/death were compared between the two drug groups in Period 1 using Cox proportional hazards models, with time since treatment as the time scale. The Cox models were stratified by STP areas to account for geographic heterogeneity in baseline hazards, with sequential adjustment for other baseline covariates. Model 1 was adjusted for age and sex; Model 2 additionally adjusted for ten high-risk cohort categories; Model 3 further adjusted for ethnicity, IMD quintiles, vaccination status, calendar week; and Model 4 additionally adjusted for BMI category, diabetes, hypertension, chronic cardiac and respiratory diseases. The proportional hazards assumption was assessed by testing for a zero slope in the scaled Schoenfeld residuals for each Cox model.

We further adopted the propensity score weighting (PSW) method as an alternative approach to account for confounding bias.[9] We used PSW to balance the distributions of relevant covariates between the two drug groups. The propensity score for each patient was defined as the conditional probability of being treated with sotrovimab, estimated with a binary logistic regression of the actual treatment allocation on relevant baseline covariates. The average treatment effect (ATE) weighting scheme was then applied to the Cox model based on the estimated propensity scores. Balance check of baseline covariates after weighting was conducted using standardised mean differences between groups (with threshold of <0.10 as the indicator of well-balanced). Robust variance estimators were used in the weighted Cox models. Missing values of covariates were treated as separate categories in the main analyses.

Similar analytical procedures were used for comparing risks of secondary outcomes between groups. In addition, we explored whether the following factors could modify the observed comparative effectiveness: each high-risk cohort, COVID-19 vaccination status (three or more vs. less than three), BMI categories (≥30 vs. <30 kg/m^2^), presence of diabetes, hypertension, chronic cardiac diseases or chronic respiratory diseases, days between test positive and treatment initiation (<3 vs. 3-5 days), age group (<60 vs. ≥60 years), sex and ethnicity (White vs. non-White). We sought to test effect modification by each of these variables by adding the corresponding interaction term between the variable and drug group in the stratified Cox model.

Additional sensitivity analyses based on the stratified Cox model were conducted to assess the robustness of the main findings, including (1) using complete case analysis or Multiple Imputation by Chained Equations for missing values (given the assumption of missing-at-random) instead of treating missing values as a separate category; (2) using Cox models stratified by calendar weeks to account for potential temporal heterogeneity in baseline hazards, with conventional adjustment for other covariates; (3) additionally adjusting for time between test positive and treatment initiation, and time between last vaccination date and treatment initiation; (4) using restricted cubic splines for age to further control for potential non-linear age effect; (5) excluding patients with treatment records of both sotrovimab and molnupiravir, or with treatment records of any other therapies (i.e., casirivimab, Paxlovid, or remdesivir); (6) excluding patients who did not have a positive SARS-CoV-2 test record before treatment or initiated treatment after 5 days since positive SARS-CoV-2 test; (7) creating a 1-day or 2-day lag in the follow-up start date to account for potential delays in drug administration (i.e., start the follow-up on the second day or the third day after the recorded treatment date).

Finally, to assess whether the main findings during Period 1 when the Omicron BA.1 was the dominant variant in England (December 2021-February 2022) [8] persist in the BA.2 era, we conducted an exploratory analysis using data from patients treated during Period 2 following similar analytical approaches.

### Software and reproducibility

Data management was performed using Python, with analysis carried out using Stata 16.1. Code for data management and analysis, as well as codelists, are archived online (https://github.com/opensafely/sotrovimab-and-molnupiravir). All iterations of the pre-specified study protocol are archived with version control (https://github.com/opensafely/sotrovimab-and-molnupiravir/tree/main/docs).

### Patient and public involvement

We have developed a publicly available website https://opensafely.org/ through which we invite any patient or member of the public to make contact regarding this study or the broader OpenSAFELY project.

## Results

### Patient characteristics

Between December 16, 2021 and February 10, 2022, a total of 6020 non-hospitalised COVID-19 patients within the OpenSAFELY-TPP platform and who met the study criteria were treated with sotrovimab (n=3331) or molnupiravir (n=2689; **Figure 1**). The mean age of the 6020 patients was 52.3 (SD=16.0) years; 58.8% were female, 88.7% were White and 87.6% had three or more COVID-19 vaccinations. Compared to those treated with molnupiravir, those treated with sotrovimab were slightly younger (mean age: 51.7 vs. 52.9 years), had a lower proportion of patients with Down syndrome (1.3% vs. 3.5%), immunosuppression (17.6% vs. 20.5%) and HIV/AIDS (2.2% vs. 4.4%). In contrast, there were a higher proportion of patients with renal disease (15.3% vs. 9.9%), solid organ transplant recipients (15.1% vs. 11.3%), and patients with obesity (36.5% vs. 34.4%) in the sotrovimab group compared to the molnupiravir group. The two groups were similar with respect to a wide range of other characteristics (**Table 1**).

### Comparative effectiveness for the primary outcome

Among the 6020 patients treated with sotrovimab or molnupiravir, 87 cases (1.45%) of COVID-19 related hospitalisations/deaths were observed during the 28 days of follow-up after treatment initiation; 32 (0.96%) in the sotrovimab group and 55 (2.05%) in the molnupiravir group. Of these 87 patients, 25 (0.42%) died of COVID-19 during the 28 days of follow-up (7 in the sotrovimab group and 18 in the molnupiravir group), among whom 16 died after COVID-19 related hospitalisation and 9 died without hospitalisation.

Results of stratified Cox regression showed that, after adjusting for demographic variables, ten high-risk cohort categories, vaccination status, calendar week, BMI category and other comorbidities, treatment with sotrovimab was associated with substantially lower risk of 28-day COVID-19 related hospitalisation/death than treatment with molnupiravir (hazard ratio, HR=0.54, 95% CI: 0.33 to 0.88; P=0.014). Consistent results favouring sotrovimab over molnupiravir were obtained from propensity score weighted Cox models (Model 4: HR=0.50, 95% CI: 0.31 to 0.81; P=0.005), following confirmation of successful balance of baseline covariates between groups in the weighted sample (**Supplementary Figure 1**). The magnitude of HRs was stable during the sequential covariate adjustment process (ranging from 0.46-0.55 across different models; **Figure 2**). No violation of the proportional hazards assumption was detected in any model (P>0.10).

**Figure 2.**
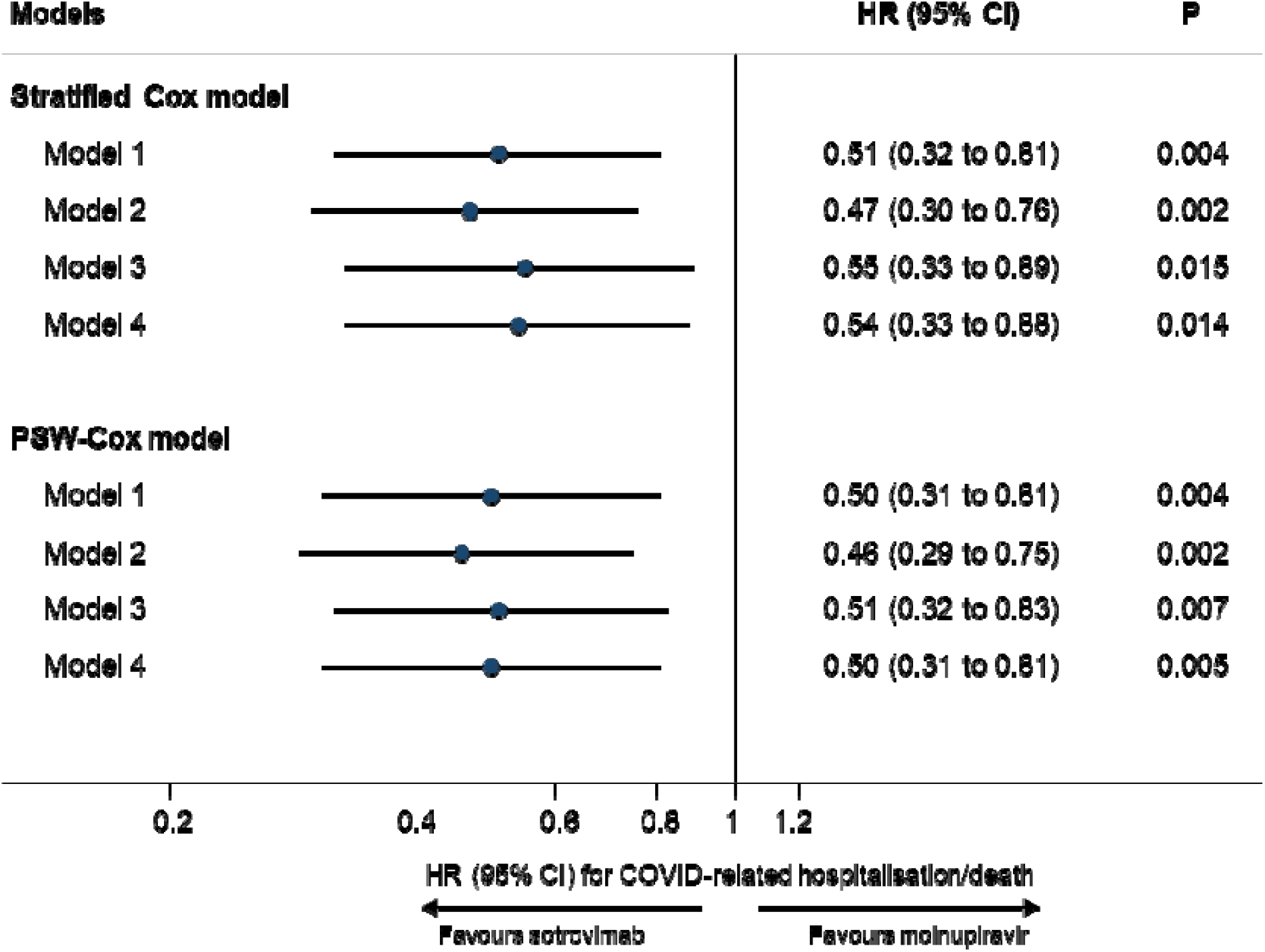
Comparing risk of 28-day COVID-19 related hospitalisation/death between patients treated with sotrovimab vs. molnupiravir. Note: HR=hazard ratio; CI=confidence interval; PSW=propensity score weighting. Model 1 adjusted for age and sex; Model 2 additionally adjusted for ten high-risk cohort categories; Model 3 further adjusted for ethnicity, IMD quintiles, vaccination status, calendar week; and Model 4 additionally adjusted for BMI category, diabetes, hypertension, chronic cardiac and respiratory diseases.

### Comparative effectiveness for secondary outcomes

For the secondary outcomes, 95 cases (1.58%) of COVID-19 related hospitalisations/deaths were observed during the 60 days of follow-up after treatment initiation (34 in the sotrovimab group and 61 in the molnupiravir group). Results of stratified Cox regression showed a significantly lower risk in the sotrovimab group than in the molnupiravir group (HRs ranging from 0.46-0.51 in Models 1-4, all P values <0.05; **Table 2**). During the 28 days of follow-up after treatment initiation, 250 cases (4.17%) of all-cause hospitalisations or deaths were observed, with 127 cases (3.83%) in the sotrovimab group and 123 cases (4.58%) in the molnupiravir group. Unlike the COVID-19 specific outcomes, no significant difference in risk of all-cause hospitalisation/death was detected between the sotrovimab group and the molnupiravir group in stratified Cox regressions (HRs ranging from 0.84-0.96 in Models 1-4, all P values >0.05; **Table 2**).

**Table 2.** Comparison of risks of secondary outcomes between patients treated with sotrovimab vs. molnupiravir.

| Secondary outcomes | N | Events | HR (95% CI) for sotrovimab (ref=molnupiravir) |
| --- | --- | --- | --- |
| 60-day COVID-19 related hospitalisation/death | 6020 | 95 |  |
| Model 1 (age-sex adjusted) |  |  | 0.50 (0.32 to 0.79) |
| Model 2 (+high-risk cohort categories adjusted) |  |  | 0.46 (0.29 to 0.73) |
| Model 3 (+demographics, vaccination and calendar time adjusted) |  |  | 0.51 (0.32 to 0.82) |
| Model 4 (+comorbidities adjusted) |  |  | 0.50 (0.31 to 0.81) |
| 28-day all-cause hospitalisation/death | 6001 | 250 |  |
| Model 1 (age-sex adjusted) |  |  | 0.96 (0.73 to 1.26) |
| Model 2 (+high-risk cohort categories adjusted) |  |  | 0.84 (0.64 to 1.10) |
| Model 3 (+demographics, vaccination and calendar time adjusted) |  |  | 0.87 (0.66 to 1.16) |
| Model 4 (+comorbidities adjusted) |  |  | 0.86 (0.65 to 1.14) |

Note: HR=hazard ratio; CI=confidence interval. Results were based on the Cox model stratified by area. Model 1 adjusted for age and sex; Model 2 additionally adjusted for ten high-risk cohort categories; Model 3 further adjusted for ethnicity, IMD quintiles, vaccination status, calendar week; and Model 4 additionally adjusted for BMI category, diabetes, hypertension, chronic cardiac and respiratory diseases.

### Sensitivity analyses and tests for effect modification

Results of sensitivity analyses were consistent with the main findings (HR for the primary outcome ranging from 0.51-0.57 across different analyses; **Table 3**). No substantial effect modification was observed for any of the ten high-risk cohort categories, COVID-19 vaccination status, presence of obesity, diabetes, hypertension, chronic cardiac diseases or chronic respiratory diseases, time since test positive, age group, sex or ethnicity (P for interaction>0.10; **Supplementary Figure 2**). Similar results to the main analysis were observed within the subset of 5271 patients who had three or more COVID-19 vaccinations (Model 4 for the primary outcome: HR=0.53 for sotrovimab vs. molnupiravir, 95% CI: 0.31 to 0.90; P=0.019).

**Table 3.** Sensitivity analyses for risk of 28-day COVID-19 related hospitalisation/death comparing patients treated with sotrovimab vs. molnupiravir.

| Sensitivity analyses | N | Events | HR (95% CI) for sotrovimab (ref=molnupiravir) |
| --- | --- | --- | --- |
| Main analysis (for comparison purpose) | 6020 | 87 | 0.54 (0.33 to 0.88) |
| Complete case analysis | 5214 | 74 | 0.53 (0.31 to 0.91) |
| Multiple imputation for missing values in covariates | 6020 | 87 | 0.54 (0.33 to 0.89) |
| Using Cox models stratified by calendar weeks | 6020 | 87 | 0.53 (0.33 to 0.88) |
| Additionally adjusting for time between test positive or last vaccination date and treatment initiation | 6020 | 87 | 0.52 (0.32 to 0.86) |
| Using restricted cubic splines to control for age effect | 6020 | 87 | 0.52 (0.32 to 0.86) |
| Excluding patients with treatment records of both sotrovimab and molnupiravir, or with any other therapies | 6010 | 87 | 0.54 (0.33 to 0.88) |
| Excluding patients without positive test record before treatment or initiated treatment after 5 days since positive test | 5725 | 82 | 0.56 (0.33 to 0.93) |
| Creating a 1-day lag in the follow-up start date | 6001 | 74 | 0.51 (0.30 to 0.87) |
| Creating a 2-day lag in the follow-up start date | 5985 | 63 | 0.57 (0.32 to 1.02) |

Note: HR=hazard ratio; CI=confidence interval. Sensitivity analyses were based on the fully-adjusted stratified Cox model (Model 4).

### Exploratory analyses of comparative effectiveness during the Omicron BA.2 era

A further 7949 COVID-19 patients treated with sotrovimab (n=5979) or molnupiravir (n=1970) between February 16 and May 1, 2022 were included in the exploratory analysis (**Figure 1**). The patients included during Period 2 were averagely older (mean age=58.8, SD=16.0) and had a higher proportion of White people (95.2%) and fully vaccinated people (94.0%) than those treated during Period 1. Compared to those treated with molnupiravir, those treated with sotrovimab were younger (mean age: 57.9 vs. 61.4 years), had a lower proportion of patients with Down syndrome (1.4% vs. 3.4%), immune-mediated inflammatory disorders (44.7% vs. 47.7%), diabetes (22.7% vs. 25.3%), chronic cardiac diseases (17.9% vs. 21.5%) and hypertension (46.9% vs. 50.8%), and had a higher proportion of solid organ transplant recipients (15.0% vs. 11.0%) and patients with haematological disease (18.3% vs. 14.9%). The two groups were similar with respect to other characteristics (**Supplementary Table 2**).

Among the 7949 patients, 97 cases (1.22%) of COVID-19 related hospitalisations/deaths were observed during the 28 days of follow-up; 57 (0.95%) in the sotrovimab group and 40 (2.03%) in the molnupiravir group. Of these 97 patients, 28 (0.35%) died of COVID-19 during the 28 days of follow-up (9 in the sotrovimab group and 19 in the molnupiravir group). Treatment with sotrovimab was associated with substantially lower risk of 28-day COVID-19 related hospitalisation/death than treatment with molnupiravir in both stratified Cox regression (Model 4: HR=0.44, 95% CI: 0.27 to 0.71; P=0.001) and propensity score weighted Cox model (Model 4: HR=0.53, 95% CI: 0.32 to 0.86; P=0.010; **Supplementary Figures 3-4**). The magnitude of HRs was stable during the sequential covariate adjustment process (ranging from 0.43-0.55 across different models; **Supplementary Figure 3**). No violation of the proportional hazards assumption was detected in any model (P>0.10).

## Discussion

### Summary

In this national, real-world cohort study we have assessed the comparative effectiveness of sotrovimab and molnupiravir in preventing severe COVID-19 outcomes among non-hospitalised COVID-19 patients. We utilised the multi-sourced electronic health record data within the OpenSAFELY-TPP platform to provide timely evidence to guide COVID-19 clinical management. We focused on patients treated between December 16, 2021 and February 10, 2022 in the main analysis to make sure the two drug groups were comparable and reduce confounding by indication based on clinical guidelines at that time.[3] The results showed a consistent and robust effect estimate of lower risk of COVID-19 related hospitalisation/death among those treated with sotrovimab compared to molnupiravir after applying different analytical approaches and when adjusting for a wide range of potential confounders, and in subgroup analysis including those underrepresented in clinical trials. The results persisted in an exploratory analysis of patients receiving treatments between February 16 and May 1, 2022, when Omicron BA.2 was the dominant variant in England.

### Findings in context

Our findings are in line with the published trial results despite the study period in the Omicron era. The COMET-ICE trial [4] was a phase 3 double-blind RCT that evaluated the use of sotrovimab in unvaccinated, non-hospitalised, high-risk adult patients with symptomatic COVID-19. An interim analysis with 583 patients from four countries showed a reduced risk of all-cause hospitalisation or death within 28 days in the sotrovimab group compared with the placebo group (1% vs. 7%, P=0.002).[4] Similar results were reported for the final sample of 1057 patients from five countries, with the risk estimate of 1% with sotrovimab vs. 6% with placebo (adjusted relative risk=0.21; absolute risk difference=-4.53%, 95% CI: -6.70% to -2.37%; P<0.001).[10] In contrast, a weaker effect was obtained from the phase 3 component of MOVe-OUT trial [5] for molnupiravir, which was also a double-blind RCT in non-hospitalised, unvaccinated adults with mild-to-moderate COVID-19 and at least one risk factor for severe illness. The interim results of 755 participants from 15 countries showed that the risk of all-cause hospitalisation or death during the 28-day follow-up was lower with molnupiravir than with placebo (7.3% vs. 14.1%; absolute risk difference=-6.8%, 95% CI: -11.3% to -2.4%; P=0.001).[5] However, in the final sample of 1433 participants from 20 countries, a lower efficacy was observed, with the risk estimate of 6.8% in the molnupiravir group vs. 9.7% in the placebo group (relative risk=0.70; absolute risk difference=-3.0%, 95% CI: -5.9% to -0.1%; P=0.043).[5]

Evidence from several *in vitro* or *in vivo* studies also suggested that both sotrovimab and molnupiravir remained active against the Omicron BA.1 variant (the predominant variant during the treatment period of our main analysis [8]).[11-14] However, there have been concerns regarding the possible loss of efficacy of sotrovimab against the Omicron BA.2 variant, though the existing evidence has been contradictory. For example, the Omicron BA.2 sublineage was shown to exhibit marked resistance to sotrovimab in *in vitro* experiments,[15,16] whereas an *in vivo* experiment found both molnupiravir and sotrovimab can restrict viral replication in the lungs of BA.2-infected hamsters.[17] Our exploratory analysis in the BA.2 era supported the persistent protective role of sotrovimab against this sub-variant, which was also in line with the preliminary epidemiological data reported by the UK Health Security Agency that the hospitalisation risk after sotrovimab use has remained similar between the periods when the BA.1 and BA.2 variants were most prevalent.[18]

### Policy implications and interpretation

The current clinical guideline from the NHS England [7] has de-prioritised molnupiravir for routine clinical use in non-hospitalised high-risk symptomatic COVID-19 patients based on the recent trial results.[5] Sotrovimab is recommended as one of the first-line treatment options (along with Paxlovid), whilst molnupiravir is considered a third-line option (following a second-line antiviral: remdesivir), only recommended when the other drugs cannot be used due to contraindications or feasibility issues. However, no comparative effectiveness trial has been conducted to support such clinical pathways. Our real-world findings within a time period when both drugs were frequently prescribed provide supportive evidence for this updated guideline. Assuming that molnupiravir had limited or no impact on COVID-19 outcomes, our results imply that sotrovimab substantially reduced the risk of COVID-19 related hospitalisation/death when comparing with eligible patients who did not receive sotrovimab or other drugs in real-world settings.

More importantly, given that both the COMET-ICE and MOVe-OUT trials only recruited unvaccinated patients, there has been uncertainty about the effects in vaccinated populations.[3,19] This is a concern for both sotrovimab (whether vaccine-induced active immunity influences passive immunisation with nMAbs [20]) and molnupiravir (given the preliminary finding of limited efficacy in seropositive patients from the MOVe-OUT trial [5]). Our analysis restricted to patients with three or more COVID-19 vaccinations supports the conclusion that sotrovimab remains beneficial in fully vaccinated patients, which now represent the majority of the COVID-19 patient population in many settings.[21]

### Strengths and weaknesses

The key strengths of this study are the scale, level of detail and completeness of the underlying primary care EHR data and the linkage to multiple COVID-19 relevant national databases within the OpenSAFELY-TPP platform. In addition, the concurrent national rollout of sotrovimab and molnupiravir under similar indications between December 16, 2021 and February 10, 2022 enables us to make direct head-to-head comparisons of their effectiveness.

Several limitations of this study need to be considered. The patients included in this study are assumed to be only those who met the eligibility criteria made by NHS England,[3] thus limiting further generalisation of our findings to people not in a known high-risk group. In addition, the possibility of residual confounding cannot be ruled out in this real-world observational study, in particular related to differences in disease severity or other unmeasured features that may have influenced clinician’s choice of therapy at assessment. This could be more evident in the exploratory analyses after February 2022, when there was no longer clinical equipoise between prescribing guidelines for sotrovimab and molnupiravir. Therefore, these findings should be interpreted with caution. However, given the size of the observed effect and its robustness across multiple sensitivity analyses, such bias would have to be substantial to fully explain the findings. Finally, our results cannot be used to infer lack of efficacy of molnupiravir for use in community settings; results from large-scale RCTs, such as the UK PANORAMIC trial (https://www.panoramictrial.org/), are needed to draw such causal conclusions.

### Future research

Despite the potential benefits of therapeutic options for non-hospitalised COVID-19 patients in preventing COVID-19 related hospitalisation/death, there are still some safety concerns to be explored using real-world data. Besides mild or moderate post-treatment symptoms reported during the trials, some uncommon side effects like urticaria and anaphylaxis have been observed for sotrovimab,[22] and a preclinical study of molnupiravir suggested a possibility of bone marrow suppression and thrombocytopenia.[23] Immediate post-marketing surveillance, especially with large-scale EHR data, is vital to comprehensively characterise and quantify the risk-benefit balance for these newly available drugs.

On the other hand, the lower baseline risk of severe outcomes [24-27] due to current prevalence of Omicron variants and high population rates of vaccination and/or prior infection, could result in lower absolute risk reduction by these medications. This situation may change with future variants, as may the effectiveness of nMAbs and antivirals. Cost-effectiveness studies of administration of nMAbs and antivirals in non-hospitalised COVID-19 patients may also be informative,[21,28] especially for nMAbs in consideration of higher price and administration costs.

### Conclusion

In conclusion, our findings suggest that in routine care sotrovimab was associated with a substantially lower risk of severe COVID-19 outcomes compared to molnupiravir for non-hospitalised high-risk COVID-19 adult patients in England, including those who were fully vaccinated. This study shows that early post-implementation monitoring of drug effects can be used to provide direct evidence to support treatment decisions, and results are consistent with current UK guidelines favouring use of sotrovimab over molnupiravir.

## Data Availability

Access to the underlying identifiable and potentially re-identifiable pseudonymised electronic health record data is tightly governed by various legislative and regulatory frameworks, and restricted by best practice. The data in OpenSAFELY is drawn from General Practice data across England where TPP is the data processor. TPP developers initiate an automated process to create pseudonymised records in the core OpenSAFELY database, which are copies of key structured data tables in the identifiable records. These pseudonymised records are linked onto key external data resources that have also been pseudonymised via SHA-512 one-way hashing of NHS numbers using a shared salt. Bennett Institute for Applied Data Science developers and PIs holding contracts with NHS England have access to the OpenSAFELY pseudonymised data tables as needed to develop the OpenSAFELY tools. These tools in turn enable researchers with OpenSAFELY data access agreements to write and execute code for data management and data analysis without direct access to the underlying raw pseudonymised patient data, and to review the outputs of this code. All code for the full data management pipeline-from raw data to completed results for this analysis-and for the OpenSAFELY platform as a whole is available for review at github.com/OpenSAFELY. Detailed pseudonymised patient data is potentially re-identifiable and therefore not shared.

## Acknowledgements

We are very grateful for all the support received from the TPP Technical Operations team throughout this work, and for generous assistance from the information governance and database teams at NHS England / NHSX.

## Contributors

LAT, BG, BZ, AJW, SJWE, IJD, and BM conceptualised and designed the study. BZ, ACAG, and JT contributed to the coding and statistical analysis. BZ and LAT drafted the original version of the manuscript. AM, BG, CB, JP, CEM, HJC, and SCJB contributed to information governance and project administration. All authors contributed to interpretation of the results and critical revision of the manuscript. All authors approved the final manuscript. LAT and BG are guarantors for this study. The corresponding author attests that all listed authors meet authorship criteria and that no others meeting the criteria have been omitted.

## Data access and verification

Access to the underlying identifiable and potentially re-identifiable pseudonymised electronic health record data is tightly governed by various legislative and regulatory frameworks, and restricted by best practice. The data in OpenSAFELY is drawn from General Practice (GP) data across England where TPP is the data processor. TPP developers initiate an automated process to create pseudonymised records in the core OpenSAFELY database, which are copies of key structured data tables in the identifiable records. These pseudonymised records are linked onto key external data resources that have also been pseudonymised via SHA-512 one-way hashing of NHS numbers using a shared salt. Bennett Institute for Applied Data Science developers and PIs holding contracts with NHS England have access to the OpenSAFELY pseudonymised data tables as needed to develop the OpenSAFELY tools. These tools in turn enable researchers with OpenSAFELY data access agreements to write and execute code for data management and data analysis without direct access to the underlying raw pseudonymised patient data, and to review the outputs of this code. All code for the full data management pipeline, from raw data to completed results for this analysis, and for the OpenSAFELY platform as a whole is available for review at github.com/OpenSAFELY.

## Information governance and ethical approval

NHS England is the data controller for OpenSAFELY-TPP; TPP is the data processor; all study authors using OpenSAFELY have the approval of NHS England. This implementation of OpenSAFELY is hosted within the TPP environment which is accredited to the ISO 27001 information security standard and is NHS IG Toolkit compliant. Patient data has been pseudonymised for analysis and linkage using industry standard cryptographic hashing techniques; all pseudonymised datasets transmitted for linkage onto OpenSAFELY are encrypted; access to the platform is via a virtual private network (VPN) connection, restricted to a small group of researchers; the researchers hold contracts with NHS England and only access the platform to initiate database queries and statistical models; all database activity is logged; only aggregate statistical outputs leave the platform environment following best practice for anonymisation of results such as statistical disclosure control for low cell counts.

The OpenSAFELY research platform adheres to the obligations of the UK General Data Protection Regulation (GDPR) and the Data Protection Act 2018. In March 2020, the Secretary of State for Health and Social Care used powers under the UK Health Service (Control of Patient Information) Regulations 2002 (COPI) to require organisations to process confidential patient information for the purposes of protecting public health, providing healthcare services to the public and monitoring and managing the COVID-19 outbreak and incidents of exposure; this sets aside the requirement for patient consent. Taken together, these provide the legal bases to link patient datasets on the OpenSAFELY platform. GP practices, from which the primary care data are obtained, are required to share relevant health information to support the public health response to the pandemic, and have been informed of the OpenSAFELY analytics platform.

This study was approved by the Health Research Authority (REC reference 20/LO/0651) and by the London School of Hygiene & Tropical Medicine’s Ethics Board (reference 21863).

## Transparency statement

The lead authors (LAT and BG) affirm that the manuscript is an honest, accurate, and transparent account of the study being reported; that no important aspects of the study have been omitted; and that any discrepancies from the study as planned (and, if relevant, registered) have been explained.

## Copyright/license for publication

The Corresponding Author has the right to grant on behalf of all authors and does grant on behalf of all authors, a worldwide licence to the Publishers and its licensees in perpetuity, in all forms, formats and media (whether known now or created in the future), to i) publish, reproduce, distribute, display and store the Contribution, ii) translate the Contribution into other languages, create adaptations, reprints, include within collections and create summaries, extracts and/or, abstracts of the Contribution, iii) create any other derivative work(s) based on the Contribution, iv) to exploit all subsidiary rights in the Contribution, v) the inclusion of electronic links from the Contribution to third party material where-ever it may be located; and, vi) licence any third party to do any or all of the above.

