## Supplementary Table 1 for "Comparative effectiveness of sotrovimab and molnupiravir for prevention of severe COVID-19 outcomes in non-hospitalised patients: an observational cohort study using the OpenSAFELY platform"

### Supplementary Materials

**Supplementary Table 1. Detected day case admissions that were not counted as hospitalisation events.**

| Admissions of which the admission and discharge dates were the same | Number of events | |
|---|---|---|
| | **Molnupiravir** | **Sotrovimab** |
| **Period 1 (main analysis)** | | |
| Detected day cases on Day 0 (i.e., the treatment start date) | 46 | 294 |
| Those with MAbs procedure on Day 0 | 9 | 267 |
| Detected day cases on Day 1 | <5 | 36 |
| Those with MAbs procedure on Day 1 | <5 | 26 |
| Detected day cases on or after Day 2 | <5 | 7 |
| Those with MAbs procedure on or after Day 2 | <5 | 6 |
| **Period 2 (exploratory analysis)** | | |
| Detected day cases on Day 0 (i.e., the treatment start date) | 44 | 317 |
| Those with MAbs procedure on Day 0 | 27 | 283 |
| Detected day cases on Day 1 | <5 | 39 |
| Those with MAbs procedure on Day 1 | <5 | 34 |
| Detected day cases on or after Day 2 | 6 | 17 |
| Those with MAbs procedure on or after Day 2 | <5 | 13 |

Note: The following admissions were not counted as hospitalisation events: (1) those recorded as "elective day case admission" or "regular admission" in the patient classification filed of SUS dataset, because such coding clearly indicates that the patient was admitted for a planned procedure or regular treatment (thus cannot represent severe COVID-19 outcomes); or (2) day cases that we additionally detected with the admission and discharge dates being the same. As shown in this table, most of those additionally detected day case admissions were for monoclonal antibody infusion procedure (i.e., admitted for receiving sotrovimab). Monoclonal antibody procedures were identified based on OPCS codes (X891, X892).

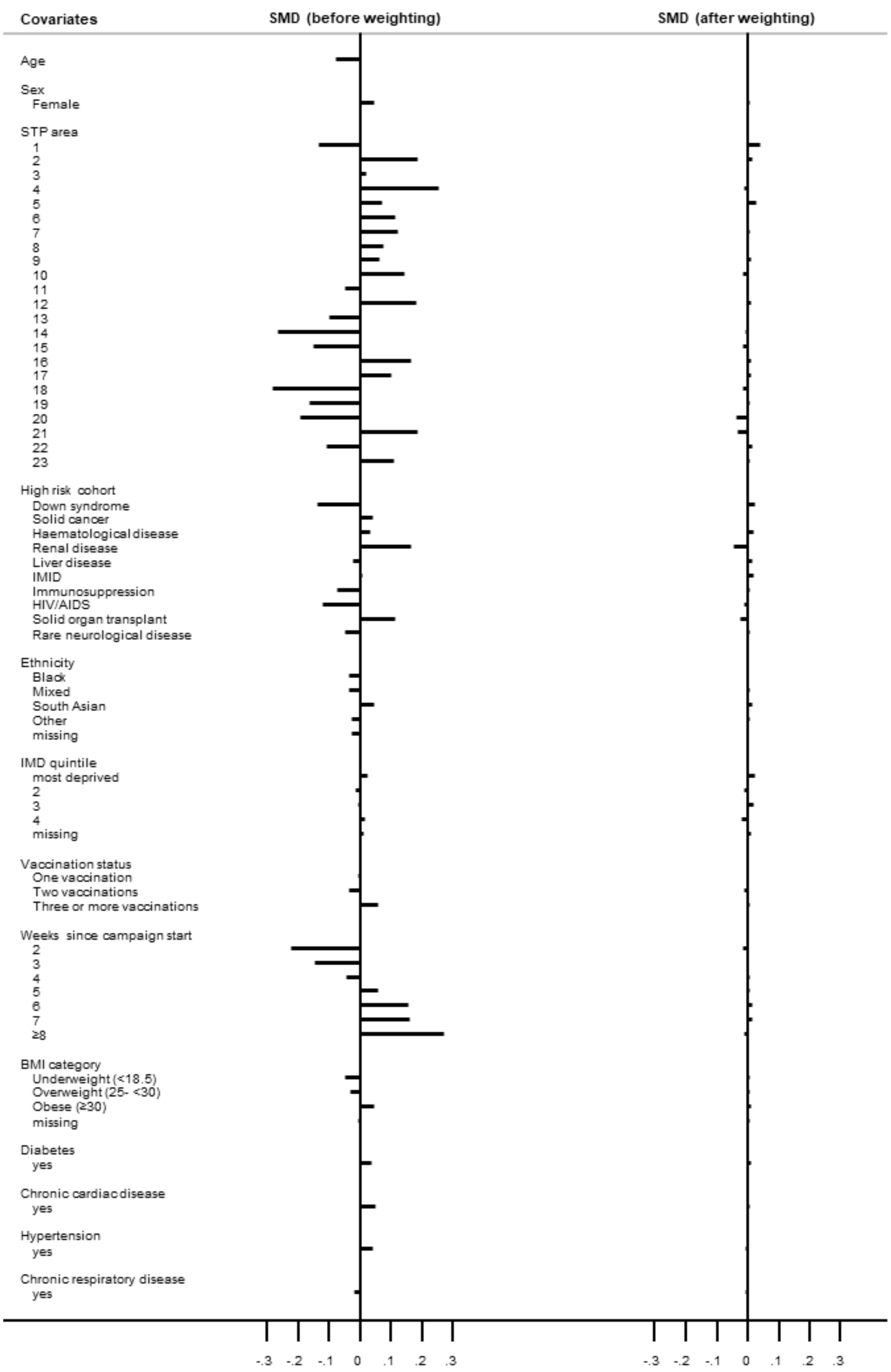


**Supplementary Figure 1. Balance check of baseline covariates between treatment groups before and after propensity score weighting (Period 1).**

Note: SMD=standardised mean difference; STP=Sustainability Transformation Partnerships; IMID=Immune-Mediated Inflammatory Diseases; IMD=Index of Multiple Deprivation; BMI=body mass index.

| Subgroups | N | Events | HR (95% CI) | $P_{interaction}$ |
|---|---|---|---|---|
| All | 6020 | 87 | 0.54 (0.33 to 0.88) | |
| Age (year) | | | | |
| <60 | 4003 | 31 | 0.32 (0.13 to 0.80) | 0.124 |
| ≥60 | 2017 | 56 | 0.57 (0.31 to 1.06) | |
| Sex | | | | |
| male | 2478 | 48 | 0.38 (0.19 to 0.78) | 0.250 |
| female | 3542 | 39 | 0.69 (0.32 to 1.48) | |
| Ethnicity | | | | |
| White | 5280 | 76 | 0.62 (0.36 to 1.04) | 0.116 |
| Non-White | 672 | 11 | 0.29 (0.02 to 4.54) | |
| Solid cancer | | | | |
| yes | 915 | 18 | 0.45 (0.11 to 1.93) | 0.489 |
| no | 5105 | 69 | 0.55 (0.31 to 0.98) | |
| Haematological disease | | | | |
| yes | 896 | 30 | 0.23 (0.08 to 0.63) | 0.500 |
| no | 5124 | 57 | 0.77 (0.41 to 1.45) | |
| Renal disease | | | | |
| yes | 775 | 18 | 0.58 (0.11 to 3.05) | 0.352 |
| no | 5245 | 69 | 0.57 (0.33 to 1.00) | |
| IMID | | | | |
| yes | 2821 | 42 | 0.74 (0.33 to 1.65) | 0.347 |
| no | 3199 | 45 | 0.41 (0.20 to 0.83) | |
| Immunosuppression | | | | |
| yes | 1138 | 23 | 0.27 (0.09 to 0.82) | 0.260 |
| no | 4882 | 64 | 0.70 (0.39 to 1.26) | |
| Solid organ transplant | | | | |
| yes | 807 | 16 | 0.28 (0.02 to 3.90) | 0.844 |
| no | 5213 | 71 | 0.50 (0.29 to 0.88) | |
| BMI (kg/m$^2$) | | | | |
| <30 | 3494 | 53 | 0.55 (0.29 to 1.05) | 0.796 |
| ≥30 | 1928 | 24 | 0.42 (0.15 to 1.18) | |
| Diabetes | | | | |
| yes | 1233 | 34 | 0.38 (0.14 to 1.02) | 0.121 |
| no | 4787 | 53 | 0.67 (0.36 to 1.25) | |
| Chronic cardiac disease | | | | |
| yes | 771 | 25 | 0.77 (0.26 to 2.27) | 0.424 |
| no | 5249 | 62 | 0.41 (0.23 to 0.75) | |
| Hypertension | | | | |
| yes | 2247 | 43 | 0.59 (0.28 to 1.25) | 0.954 |
| no | 3773 | 44 | 0.46 (0.22 to 0.93) | |
| Chronic respiratory disease | | | | |
| yes | 1199 | 42 | 0.50 (0.23 to 1.08) | 0.829 |
| no | 4821 | 45 | 0.63 (0.31 to 1.27) | |
| Days between test positive and treatment | | | | |
| ≤2 days | 3053 | 33 | 0.56 (0.24 to 1.27) | 0.205 |
| 3-5 days | 2672 | 49 | 0.51 (0.25 to 1.02) | |

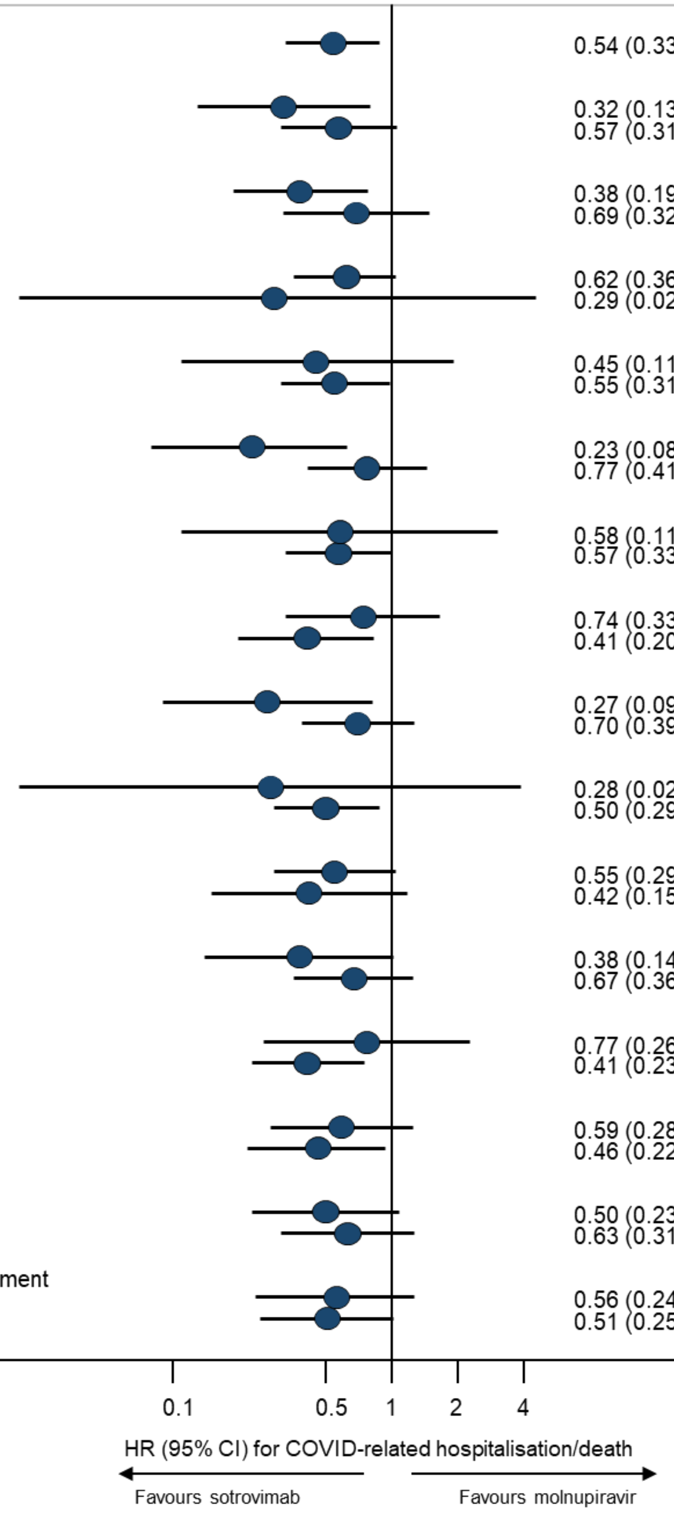

**Supplementary Figure 2. Subgroup analysis of sotrovimab vs. molnupiravir in association with risk of 28-day COVID-19 related hospitalisation/death.**

Note: HR=hazard ratio; CI=confidence interval; IMID=Immune-Mediated Inflammatory Diseases; BMI=body mass index. Subgroup analyses were based on the fully-adjusted stratified Cox model (Model 4). P for interaction between drug group and each of the following variables was: Down syndrome (0.221), liver disease (0.953), HIV/AIDS (0.999), rare neurological disease (0.999), vaccination status (0.574), respectively; no analyses within each level of these variables were done because of lack of sample size or outcome events within the subset of population.

**Supplementary Table 2. Baseline characteristics of COVID-19 patients receiving molnupiravir or sotrovimab (Period 2).**

| Characteristics | Molnupiravir group | Sotrovimab group | Total |
|---|---|---|---|
| N | 1970 | 5979 | 7949 |
| Age (year), mean (SD)* | 61.4 (16.6) | 57.9 (15.6) | 58.8 (16.0) |
| Female, n (%) | 1112 (56.5) | 3494 (58.4) | 4606 (57.9) |
| White, n (%) | 1863 (95.9) | 5632 (95.0) | 7495 (95.2) |
| Most deprived, n (%) | 213 (11.0) | 722 (12.4) | 935 (12.0) |
| Region (NHS), n (%)* | | | |
| East | 823 (41.8) | 1885 (31.5) | 2708 (34.1) |
| London | 67 (3.4) | 205 (3.4) | 272 (3.4) |
| East Midlands | 108 (5.5) | 1355 (22.7) | 1463 (18.4) |
| West Midlands | 20 (1.0) | 252 (4.2) | 272 (3.4) |
| North East | 17 (0.9) | 240 (4.0) | 257 (3.2) |
| North West | 71 (3.6) | 553 (9.2) | 624 (7.9) |
| South East | 149 (7.6) | 360 (6.0) | 509 (6.4) |
| South West | 471 (23.9) | 819 (13.7) | 1290 (16.2) |
| Yorkshire | 244 (12.4) | 310 (5.2) | 554 (7.0) |
| High-risk cohorts, n (%) | | | |
| Down syndrome* | 66 (3.4) | 84 (1.4) | 150 (1.9) |
| Solid cancer | 378 (19.2) | 1071 (17.9) | 1449 (18.2) |
| Haematological disease* | 294 (14.9) | 1094 (18.3) | 1388 (17.5) |
| Renal disease | 276 (14.0) | 901 (15.1) | 1177 (14.8) |
| Liver disease | 105 (5.3) | 323 (5.4) | 428 (5.4) |
| Immune-mediated inflammatory diseases* | 940 (47.7) | 2673 (44.7) | 3613 (45.5) |
| Immunosuppression | 308 (15.6) | 948 (15.9) | 1256 (15.8) |
| HIV/AIDS | 41 (2.1) | 101 (1.7) | 142 (1.8) |
| Solid organ transplant* | 216 (11.0) | 896 (15.0) | 1112 (14.0) |
| Rare neurological disease | 237 (12.0) | 742 (12.4) | 979 (12.3) |
| BMI (kg/$m^2$), mean (SD) | 28.8 (6.3) | 28.9 (6.6) | 28.9 (6.5) |
| Diabetes, n (%)* | 498 (25.3) | 1359 (22.7) | 1857 (23.4) |
| Chronic cardiac disease, n (%)* | 424 (21.5) | 1068 (17.9) | 1492 (18.8) |
| Hypertension, n (%)* | 1001 (50.8) | 2806 (46.9) | 3807 (47.9) |
| Chronic respiratory disease, n (%) | 493 (25.0) | 1394 (23.3) | 1887 (23.7) |
| Vaccination status, n (%) | | | |

| Characteristics | Molnupiravir group | Sotrovimab group | Total |
|---|---|---|---|
| None | 27 (1.4) | 74 (1.2) | 101 (1.3) |
| One vaccination | 15 (0.8) | 54 (0.9) | 69 (0.9) |
| Two vaccinations | 84 (4.3) | 223 (3.7) | 307 (3.9) |
| Three or more vaccinations | 1844 (93.6) | 5628 (94.1) | 7472 (94.0) |
| Days between test positive and treatment, median (IQR)* | 2 (1-3) | 2 (1-3) | 2 (1-3) |
| Weeks between campaign start and treatment, median (IQR)* | 15 (13-17) | 15 (13-17) | 15 (13-17) |

Note: SD=standard deviation; BMI=body mass index; IQR=interquartile range. Comparisons between groups were conducted using *t*-test, chi-square test or rank-sum test, where appropriate; * indicates $P<0.05$. Ethnicity, Index of Multiple Deprivation, BMI and positive test date had 79, 188, 633 and 982 missing values, respectively.

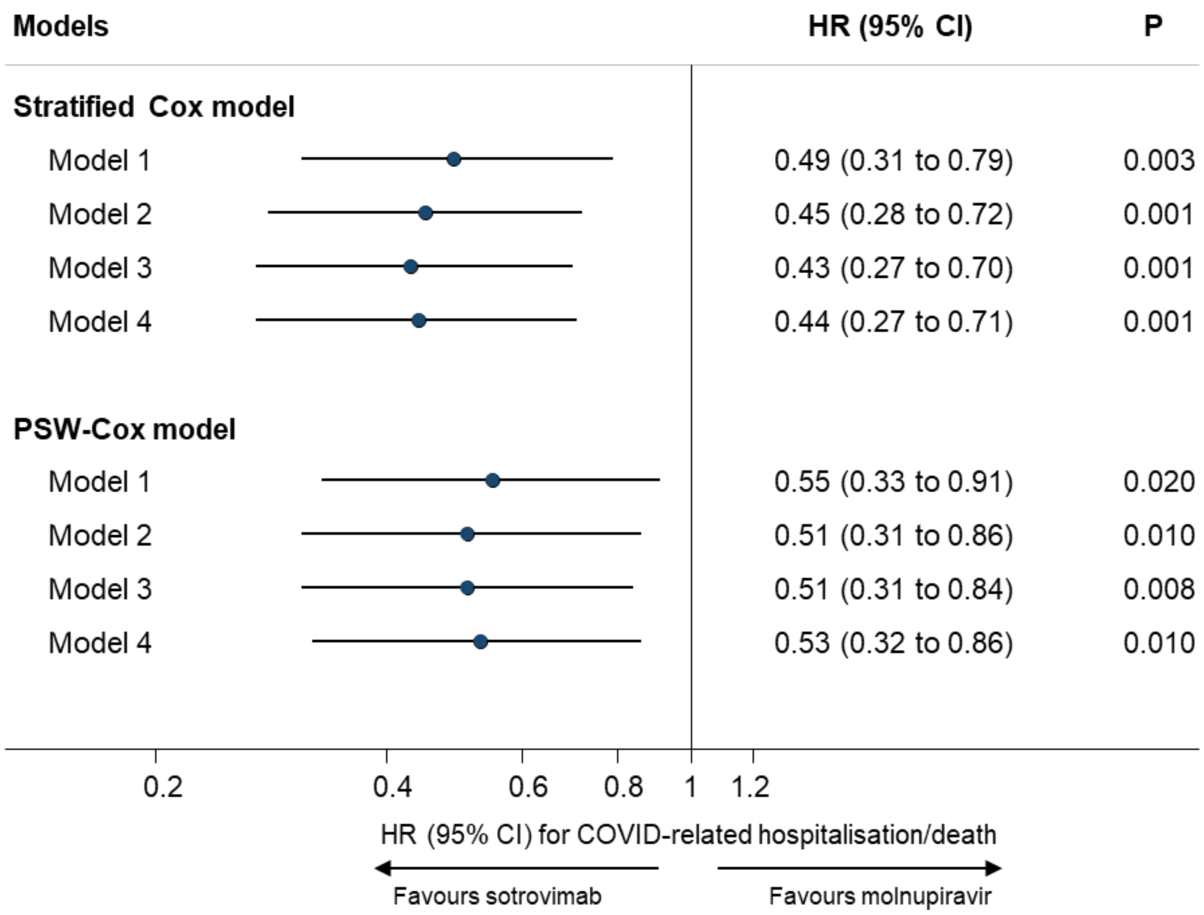


**Supplementary Figure 3. Comparing risk of 28-day COVID-19 related hospitalisation/death between patients treated with sotrovimab vs. molnupiravir (Period 2).**

Note: HR=hazard ratio; CI=confidence interval; PSW=propensity score weighting. Model 1 adjusted for age and sex; Model 2 additionally adjusted for ten high-risk cohort categories; Model 3 further adjusted for ethnicity, IMD quintiles, vaccination status, calendar week; and Model 4 additionally adjusted for BMI category, diabetes, hypertension, chronic cardiac and respiratory diseases.

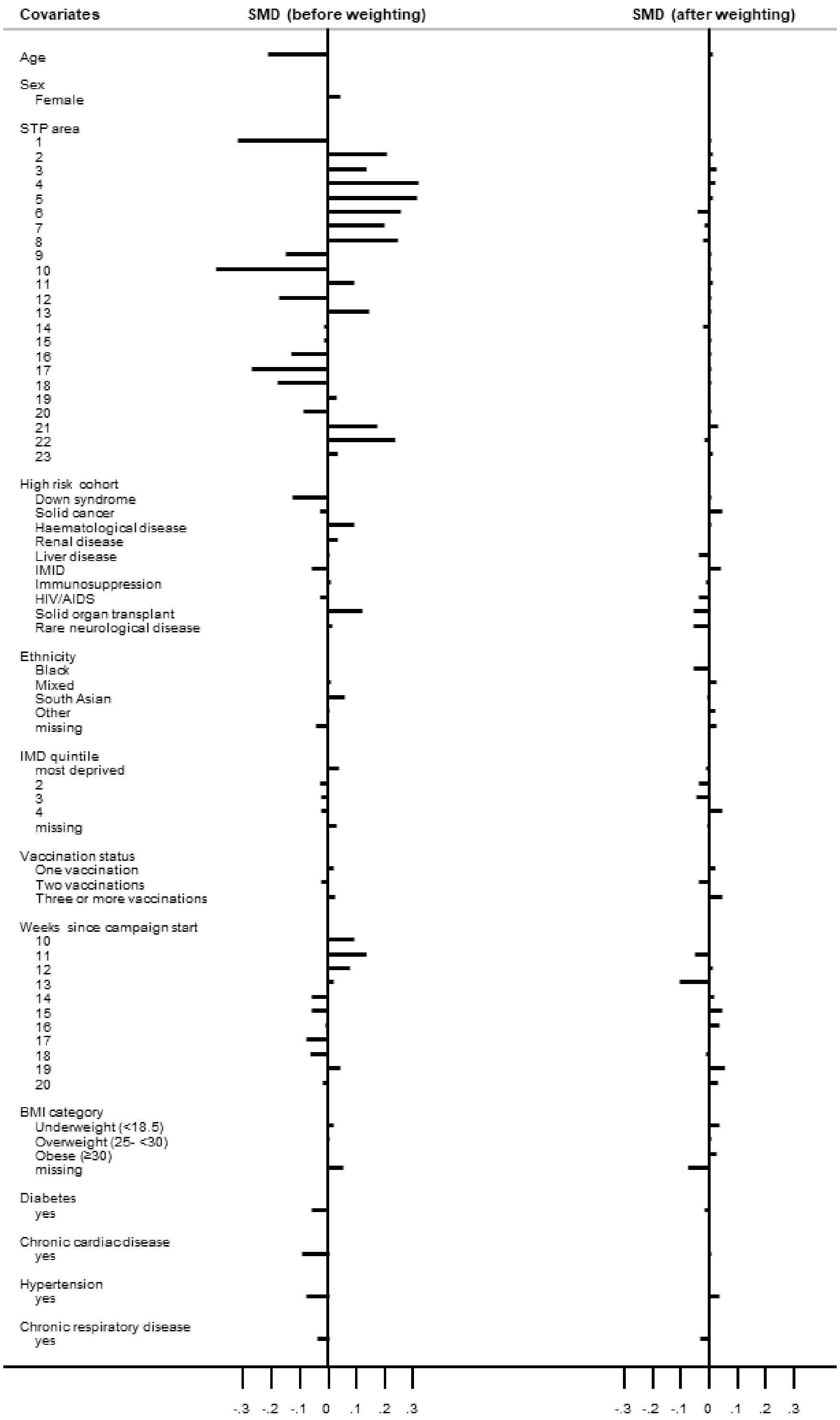


**Supplementary Figure 4. Balance check of baseline covariates between treatment groups before and after propensity score weighting (Period 2).**

Note: SMD=standardised mean difference; STP=Sustainability Transformation Partnerships; IMID=Immune-Mediated Inflammatory Diseases; IMD=Index of Multiple Deprivation; BMI=body mass index.
